# Preferences for tongue swab-versus sputum-based testing in the context of TB care: a Best-Worst Scaling exercise in Vietnam and Zambia

**DOI:** 10.1101/2024.12.22.24319450

**Authors:** Maria del Mar Castro, Hien Le, Seke Muzazu, Nam Pham, Trang Trinh, Herbert Nyirenda, Patricia Shabalu, Nora West, Ha Phan, Adithya Cattamanchi, Claudia M. Denkinger, Monde Muyoyeta, Andrew D. Kerkhoff

## Abstract

**Background:** The development of non-sputum-based tests is an urgent priority to increase access to tuberculosis (TB) diagnostic testing. Understanding preferences of people undergoing testing is critical for designing tests and strategies aligned with their needs.

**Methods:** We conducted a survey and Best-Worst Scaling (BWS) exercise to determine relative preferences for tongue swab-vs. sputum-based testing among people (≥13 years) with presumptive TB at primary health centers in Vietnam and Zambia. The BWS assessed sixteen TB test features, including accuracy, sample type, turnaround time, cost, and service aspects. We analyzed preferences using Hierarchical Bayes modeling and identified distinct preference groups using Latent Class Multinomial Logit analyses (LCA).

**Results:** Among 409 participants enrolled, 356 (87%) met quality criteria for analysis. When asked directly, most participants preferred providing tongue swabs over sputum (58% vs 29%, p<0.001; 13% no preference). In the BWS exercise, tongue swab was also preferred over sputum (mean rescaled preference weight [MPW] 6.4 [95%CI: 5.9-6.8] vs. 5.0 [95%CI: 4.6-5.4]). However, support and counseling (MPW=14.0), sensitivity (MPW=12.3), specificity (MPW=10.2), and provider attitude (MPW=7.4) were the most important features overall. Less important features included facility opening hours (MPW=3.4) and the influence of trusted sources on testing decisions (MPW=2.2). LCA identified five distinct preference groups, but support and counseling were universally valued. Participants in Groups 2 (27.3%; n=97) and 3 (17.1%; n=61) valued tongue swabs over many other features. Group 5 participants (11%; n=39) strongly valued sputum-based testing.

**Conclusions:** Participants in Vietnam and Zambia preferred tongue swab-based TB testing over sputum. However, sample type was less important than test accuracy and other TB care features affecting the testing experience.

## Introduction

Tuberculosis (TB) remains a global health challenge, contributing significantly to morbidity and mortality worldwide. The burden of TB is unevenly distributed, disproportionately affecting vulnerable populations such as individuals living in poverty, people living with HIV (PLHIV), migrants, refugees, people deprived of their liberty, and children [1]. These groups often face barriers to accessing timely and effective TB diagnosis and treatment, exacerbating health disparities and hindering global efforts to eliminate the disease [2, 3].

The development of non-sputum-based tests is an urgent priority to increase access to TB diagnostic testing. Conventional sputum-based methods are limited by their reliance on patients’ ability to produce sputum, which can be challenging for certain populations, including PLHIV and young children [4]. In addition, sputum requires complex sample processing, which increases test costs and leads to longer turnaround times [5]. The reliance on and limitations of sputum-based tests cause delays in diagnosis and treatment, which in turn contribute to ongoing transmission and poor health outcomes [3, 6, 7].

Tongue swab sampling has emerged as a promising alternative for TB, particularly for people who are unable to produce sputum [8]. Tongue swabs are easy to collect, including from PLHIV, people who are severely ill, and children [9, 10]. Recent studies have demonstrated the feasibility of swab-based sampling for TB [11]. Although diagnostic accuracy has varied and been less than that of sputum-based testing [9, 11–13], a more accessible sample such as tongue swabs can yield a similar or higher number of people diagnosed with TB, as demonstrated in a recent meta-analysis of urine lipoarabinomannan testing in hospitalized people living with HIV [5].

In addition to sample type, other features of the TB diagnosis process may be equally or more important to people affected by TB [14]. Aspects related to quality of care, such as respect and communication from the providers, are important since negative healthcare experiences reported across LMICs often stem from a lack of these elements and affect areas far beyond TB [15]. The framework developed by the Lancet Global Health Commission to measure and improve the quality of TB care emphasizes that expanding diagnosis and treatment coverage alone is insufficient to eradicate TB and that high-quality health systems are essential. Thus, values like people-centeredness [16], equity, resilience, and efficiency are critical to improve care [6] and avoid excess deaths due to poor-quality services [17].

Eliciting the preferences of people affected by TB through stated preference methods such as Best-Worst Scaling (BWS) can provide valuable insights into the features that most strongly influence acceptance and widespread adoption of new diagnostic tools and approaches [18, 19].

Understanding people’s preferences can optimize TB test development efforts and improve TB care engagement and overall TB outcomes by making diagnostic processes more accessible and acceptable to those most in need [19]. In this study, we determined the relative preferences for tongue swab-versus sputum-based molecular testing, and how they compared to other features related to TB service delivery and care in Vietnam and Zambia.

## Methods

### Study design, setting and participants

We conducted a cross-sectional survey and Best-Worst Scaling (BWS) exercise assessing different TB test and care features in Vietnam and Zambia. This evaluation was nested in a pragmatic study assessing the yield of tongue swab-versus sputum-based molecular testing in high TB burden settings. We enrolled consecutive people ≥13 years of age presenting to primary health centers in Vietnam and Zambia who met local TB testing requirements (*i.e*., a person with presumptive TB).

We excluded people unwilling or unable to provide informed consent or cognitively unable to complete the BWS based on their performance in a handbook-guided warm-up task.

### Best-Worst Scaling (BWS) Design

The BWS assessed 16 TB test features, including aspects related to sample type, cost, accuracy, service delivery, and care. These features were selected by reviewing existing literature and collaborative discussions with local investigators to maximize relevance and cultural appropriateness. **Table 1** presents the list of features, and the wording used to describe each feature in the BWS.

**Table 1.** List of features and text utilized for the BWS

| Feature | Description in the BWS |
| --- | --- |
| **Test sample collection** |  |
| **Tongue swab** | The test uses a tongue swab sample (you must stick our tongue so that it can be gently swabbed for 15 seconds) |
| **Sputum sample** | The test uses a sputum sample (you must cough thick mucus from the back of your throat into a cup) |

| **Test accuracy** |  |
| --- | --- |
| **Sensitivity (false negative)** | When you DO have TB, the chance is very low that the test gives you an incorrect negative result (the test is very good at detecting TB when you have TB) |
| **Specificity (false positive)** | When you DON'T have TB, the chance is very low that you receive unnecessary TB treatment for 6 months because the test gives you an incorrect positive result |
| **TB testing process and waiting time for results** |  |
| **Additional tests** | No additional samples or tests are needed after the first test to confirm TB diagnosis (you would not need to come back another day for further testing) |
| **Rapid results (30 minutes)** | The results of the TB test are available rapidly, on the spot (for example, within 30 minutes) |
| **Same day results (5hours)** | The results of the TB test are available the same day (for example, within 5 hours) |
| **Waiting time at facility** | You spend very little time waiting at the facility to get tested for TB (for example, less than 30 minutes) |
| **Cost and accessibility** |  |
| **Free** | The test does not cost you anything (it is free) |
| **Provider attitude** | The healthcare worker who helps you with TB testing is kind and respectful |
| **Extended opening hours** | Convenient, extended opening hours are available at the facility for TB testing (for example, early morning, nights or weekends) |
| **Community location** | Convenient community locations are available for TB testing (for example, sites near home or work) |
| **Counselling and privacy** |  |
| **Support and counseling** | You are well counseled about what will happen while taking the test, and once the results are available, you receive counseling and support on their meaning and what will happen next |
| **Privacy and stigma** | You can get tested for TB privately, without being seen by people who know you (for example, friends, colleagues, or family) |
| **Trusted source** | A trusted family member, friend, or community leader recommends you get tested for TB |
| **Results notification** | You can choose how the test results are returned to you (for example, in person at the facility, SMS or phone call) |

The BWS was designed using Lighthouse Studio software version 9.15.0 (Sawtooth software, USA) and utilized a near-Balanced Incomplete Block Design (BIBD). In a BIBD configuration (v, b, r, k, λ), ’v’ represents the number of items displayed across ’b’ questions, each containing ’k’ items. This setup achieves approximate one-way level balance with each item appearing ’r’ times, and two-way level balance as each item pairs with every other item ’λ’ times [20]. This design ensured that each participant completed one of 500 random versions of the survey, each containing 12 questions (i.e., choice tasks) that presented four features. All participants saw each feature at least 3 times, allowing for robust individual-level preference weight estimates. A single prohibition was implemented such that the two time-related features—same-day and rapid results—were not featured in the same question since the directionality of preference could be assumed. To understand the absolute importance of the 16 features, we used the dual-response indirect anchoring method [20], where each choice task was followed by a question that asked participants to indicate the importance (“all, some, none are important”) of the four displayed features (**Figure 1**). Following the BWS exercise, participants were asked two questions to assess the perceived difficulty of completing the BWS and making choices in each task. A participant information booklet was developed to provide standardized explanations to the study participants about each of the features. All informational material were delivered, and data were collected in the participants’ preferred languages (Vietnam - Vietnamese; Zambia - Nyanja, Bemba, or English).

**Figure 1.**
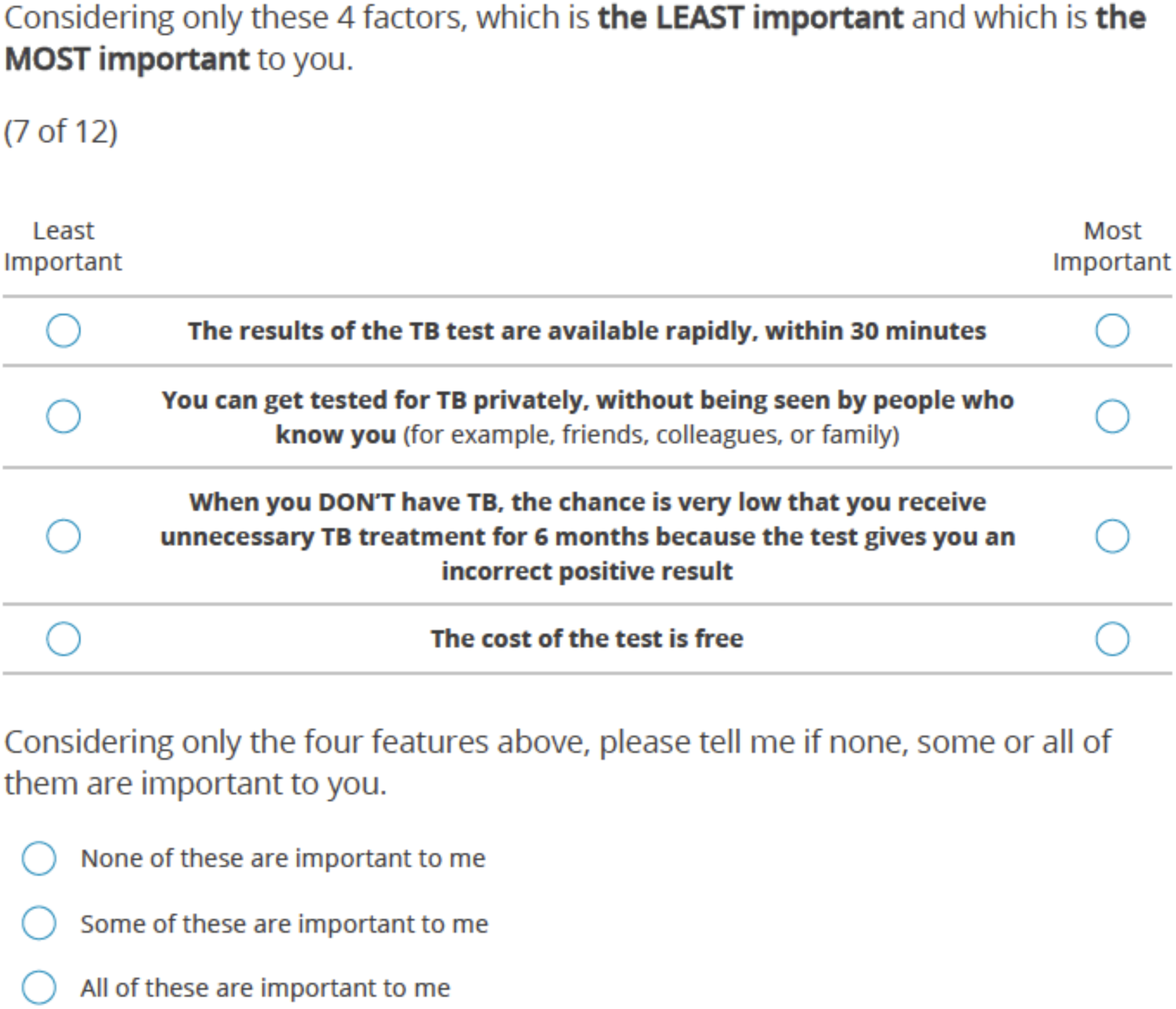
Example of a BWS choice task

To optimize the understanding and relevance of the BWS, a pilot test was initially conducted at each study site (n=4 participants in Vietnam, n=6 in Zambia). Feedback from the pilot was captured using a structured form that included aspects like time, layout of the BWS and participant information handbook, clarity, and difficulty of understanding, as well as open ended feedback for improvement. This information was used to refine the BWS, handbook and improve the quality of the translations.

### Procedures

During site visits, and based on recommendations from on-site health workers, trained study staff approached all individuals presenting to the study health centers who met the local criteria for presumptive TB and were referred for sputum-based TB testing. Prior to consenting procedures for this sub-study, all participants attempted sputum production and underwent healthcare worker-led tongue swab collection for TB testing. After informed consent for sub-study participation, trained study staff provided standardized explanations of the 16 TB testing features using a participant information booklet. Study staff then administered a brief survey capturing demographics and information related to TB testing experience including ease of sample production, discomfort, satisfaction, and direct preference for sputum vs. tongue swab collection. The BWS was administered after the survey. All data was collected using the Sawtooth Software Offline Surveys App.

### Sample size and sampling

Assuming 16 total features, where each participant is shown 4 features per task and 12 choice tasks, a minimum of 133 participants per country were required to achieve adequate statistical precision around BWS estimates [21]. We increased the target sample size to 200 per country (n=400 total) based on available budget and the study timeline.

### Statistical analysis

Prior to analysis, BWS data were assessed at the participant level to ensure quality responses. Any participant meeting two of the following three criteria had their data excluded from the analysis: (1) an individual Root Likelihood (RLH) less than the 95% RLH cut-off generated from 500 random responses in Sawtooth (suggesting they likely answered at random without understanding or evaluating the options in each task)[20]; (2) a BWS completion time less than 40% of the median overall time for respondents in that country (suggesting they rushed through each question without careful consideration of the options in each task); (3) or a self-response indicating that the BWS exercise was somewhat or very difficult to complete (suggesting they may not have fully comprehended the exercise).

We summarized participants’ baseline characteristics and direct preference survey responses descriptively. We compared differences in proportions with Fisher’s exact test. We performed Hierarchical Bayesian (HB) modeling to estimate the individual-level probability rescaled preference weights for each participant and included country as a covariate. We estimated mean rescaled preference weights (MPWs) and their 95% confidence intervals for each TB testing feature overall and by country. The rescaled MPWs, which sum to 100, indicate the relative importance of TB testing features and allow for direct comparison (*e.g*., feature X is twice as important as feature Y) since they are on the same scale.

We used Latent class multinomial logit analysis (LCA) to characterize preference heterogeneity by identifying segments of participants (groups) that cluster according to distinct preferences. For the final model, the number of distinct preference groups was selected based on optimizing statistical fit (information criterion), interpretability, group size (ensuring meaningful and reliable groups), and maximum membership probability compared to other solutions [22, 23]. HB and latent class analysis were conducted using Lighthouse software version 9.15.0 (as part of Sawtooth software).

### Ethics statement

The study was approved by the Research Ethics Committee of the Hanoi Lung Hospital in Vietnam [# 935/BVPHN-HĐĐĐ], the University of Zambia Biomedical Research Ethics Committee [# 4041-2023], the National Health Research Authority-Zambia [# NHREB002/15/07/2023], and the Ethics Committee of the Heidelberg University Hospital [#S-519/2023]. All adult participants provided informed consent prior to the survey in their preferred language. For those under 18 years of age, informed consent of the legal guardian/caretaker and assent was obtained from participants.

## Results

### Participant characteristics

Between September 2023 and February 2024, 423 eligible people were invited to participate in Vietnam and Zambia, of whom 409 completed the survey and BWS exercise. Based on the BWS quality criteria, 43 were excluded from Zambia and 10 from Vietnam, leaving 356 participants included in the final analysis (**Figure 2**). Excluded participants were younger, mainly from Zambia and more likely to report being unemployed (**Supplementary Table 1**).

**Figure 2.**
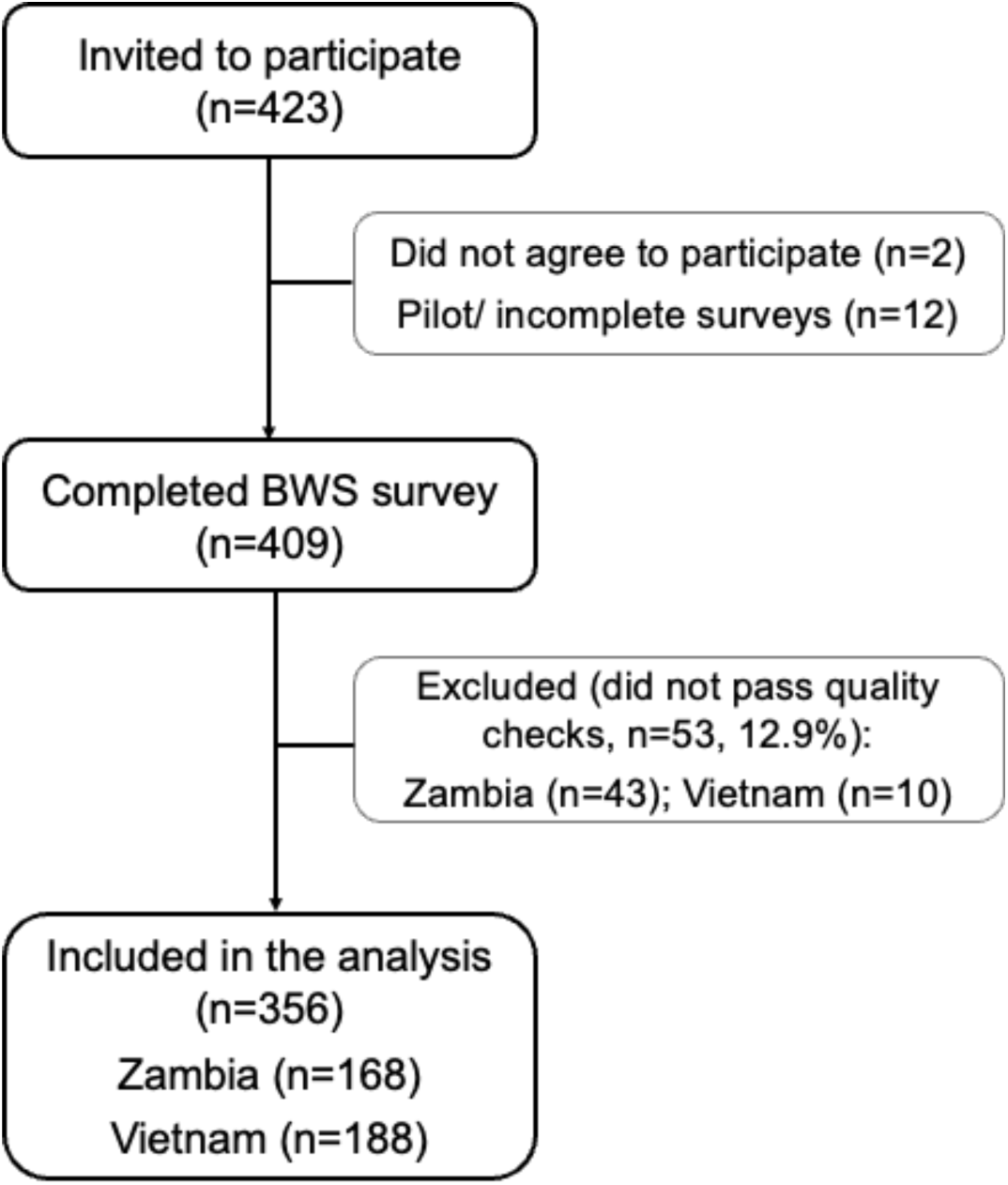
Participant enrolment and inclusion

Overall, participants had a median age of 39 years (IQR: 29-47) and the majority were female (60.7%, **Table 2**). Most participants had at least some secondary-level education and were either self-employed (30.6%) or unemployed (21.1%); 45.5% of participants had previously been tested for TB, 14.3% had received prior treatment for TB, 25% self-reported having a positive HIV status and 2.2% self-reported having diabetes. When comparing characteristics across countries, people from Vietnam were more likely to be older, have higher education status, be employed, to have been previously tested for TB, and to self-report being HIV-positive (**Table 2**).

**Table 2.** Participant demographic and clinical characteristics.

| Characteristic | Overall<br>n=356 | Vietnam<br>n=188 | Zambia<br>n=168 |
| --- | --- | --- | --- |
| **Age**, years, Median (IQR) | 39 (29-47) | 42 (37-49) | 32 (25-43) |
| **Female sex**, n (%) | 216 (60.7) | 109 (58.0) | 107 (63.7) |
| **Education**, n (%) |  |  |  |
| Never attended school | 8 (2.2) | 0 (0.0) | 8 (4.8) |
| Primary school | 63 (17.7) | 7 (3.7) | 56 (33.3) |
| Secondary school | 188 (52.8) | 97 (51.6) | 91 (54.2) |
| Higher level | 97 (27.2) | 84 (44.7) | 13 (7.7) |

| **Employment, n (%)** |  |  |  |
| --- | --- | --- | --- |
| Not employed | 75 (21.1) | 18 (9.6) | 57 (33.9) |
| Yes, informal work | 68 (19.1) | 34 (18.1) | 34 (20.2) |
| Yes, office job | 58 (16.3) | 41 (21.8) | 17 (10.1) |
| Yes, self employed | 109 (30.6) | 63 (33.5) | 46 (27.4) |
| Other | 27 (7.6) | 26 (13.8) | 1 (0.6) |
| Student | 19 (5.3) | 6 (3.2) | 13 (7.7) |
| **Prior testing for TB, n (%)** |  |  |  |
| Yes | 162 (45.5) | 93 (49.5) | 69 (41.1) |
| No | 189 (53.1) | 90 (47.9) | 99 (58.9) |
| Not sure | 5 (1.4) | 5 (2.7) | 0 (0.0) |
| **Prior TB treatment, n (%)** |  |  |  |
| Yes | 51 (14.3) | 30 (16.0) | 21 (12.5) |
| No | 305 (85.7) | 158 (84.0) | 147 (87.5) |
| **HIV status<sup>\*,#</sup>, n (%)** |  |  |  |
| Positive | 89 (25.0) | 54 (28.7) | 35 (20.8) |
| Negative | 168 (47.2) | 49 (26.1) | 119 (70.8) |
| Not sure | 98 (27.5) | 85 (45.2) | 13 (7.7) |
| **Diabetes<sup>\*</sup>, n (%)** |  |  |  |
| Yes | 8 (2.2) | 6 (3.2) | 2 (1.2) |
| No | 343 (96.3) | 177 (94.1) | 166 (98.8) |
| Not sure | 5 (1.4) | 5 (2.7) | 0 (0.0) |

\*Self-reported; #One participant declined to answer

### Sample collection and sample type preference survey

Nearly all participants were able to provide both sputum and a tongue swab sample; only three (0.8%) were unable to provide sputum, all from the Zambia site. When asked directly, most participants preferred providing tongue swabs over sputum (58.1% vs 28.7%, p<0.001), while 12.4% reported no preference; a similar proportion favored tongue swab collection across both countries (**Table 3**). Compared to sputum, tongue swabs were reported as the easier sample to collect by 74.4% of participants and were associated with lower self-reported discomfort (10.1% vs. 50.8%, p<0.001) and higher satisfaction (71.1% vs. 49.2% very satisfied, p<0.001). These trends were consistent in both country-level analyses conducted in Vietnam and Zambia (**Table 3**).

**Table 3.** Direct preferences between tongue swab and sputum testing, overall and per country

| Variable | Overall | Vietnam | Zambia | *p-value* |
| --- | --- | --- | --- | --- |
|  | n=356 | n=188 | n=168 |  |
| **Tongue swab sample** |  |  |  |  |
| Discomfort or difficulty providing the sample, n (%) | 36 (10.1) | 8 (4.3) | 28 (16.7) | <0.001 |
| Overall satisfaction with providing the sample, n (%) |  |  |  |  |
| Very satisfied | 253 (71.1) | 161 (85.6) | 92 (54.8) | <0.001 |
| Somewhat satisfied | 88 (24.7) | 25 (13.3) | 63 (37.5) |  |
| A little satisfied | 14 (3.9) | 1 (0.5) | 13 (7.7) |  |
| Not satisfied at all | 1 (0.3) | 1 (0.5) | 0 (0.0) |  |
| **Sputum sample** |  |  |  |  |
| Discomfort or difficulty providing the sample, n (%) |  |  |  |  |
| Yes | 181 (50.8) | 66 (35.1) | 115 (68.5) | <0.001 |
| No | 172 (48.3) | 122 (64.9) | 50 (29.8) |  |
| Not applicable | 3 (0.8) | 0 (0.0) | 3 (1.8) |  |
| Overall satisfaction with providing the sample, n (%) |  |  |  |  |
| Very satisfied | 175 (49.2) | 84 (44.7) | 91 (54.2) | 0.06 |
| Somewhat satisfied | 109 (30.6) | 68 (36.2) | 41 (24.4) |  |
| A little satisfied | 60 (16.9) | 32 (17.0) | 28 (16.7) |  |
| Not satisfied at all | 9 (2.5) | 4 (2.1) | 5 (3.0) |  |
| Not applicable | 3 (0.8) | 0 (0.0) | 3 (1.8) |  |
| **Preference regarding samples** |  |  |  |  |
| Easier sample to provide, n (%) |  |  |  |  |
| Sputum | 37 (10.4) | 19 (10.1) | 18 (10.7) | 0.32 |
| Tongue swab | 265 (74.4) | 141 (75.0) | 124 (73.8) |  |
| No difference | 51 (14.3) | 28 (14.9) | 23 (13.7) |  |
| No data /not applicable | 3 (0.8) | 0 (0.0) | 3 (1.8) |  |
| Preferred sample to provide for TB diagnosis, n (%) |  |  |  |  |
| Sputum | 102 (28.7) | 41 (21.8) | 61 (36.3) | <0.001 |
| Tongue swab | 207 (58.1) | 112 (59.6) | 95 (56.5) |  |
| No preference | 44 (12.4) | 35 (18.6) | 9 (5.4) |  |
| No data /not applicable | 3 (0.8) | 0 (0.0) | 3 (1.8) |  |

### Preferences for TB testing

All features assessed in this BWS were above the anchoring point, indicating that all were considered important by participants (**Supplementary Table 2**). The most important features to participants were good support and counseling (MPW=14.0 [95%CI: 13.6-14.4]), high sensitivity (MPW=12.3 [95%CI: 11.8-12.8]), high specificity (MPW=10.2 [95%CI: 9.7-10.8]), and a kind and respectful provider (MPW=7.4 [95%CI: 7.0-7.8]) (**Figure 3A**). Participants also valued not needing to undertake additional tests, a community testing location, free testing, and a choice for how they receive testing results - each had an MPW between 5.0 and 5.9. Less important features were short waiting times (MPW=3.5 [95%CI: 3.2-3.8]), extended facility hours for testing (MPW=3.4 [95%CI: 3.1-3.7]), and the influence of trusted sources on testing decisions (MPW=2.2 [95%CI: 1.9-2.5]). In terms of sample type, tongue swab was preferred over sputum (MPW=6.4 [95%CI: 5.9-6.8] vs. 5.0 [95%CI: 4.6-5.4]), with a relative rank of 5 vs. 10 out of 16, respectively (**Figure 3A**).

**Figure 3.**
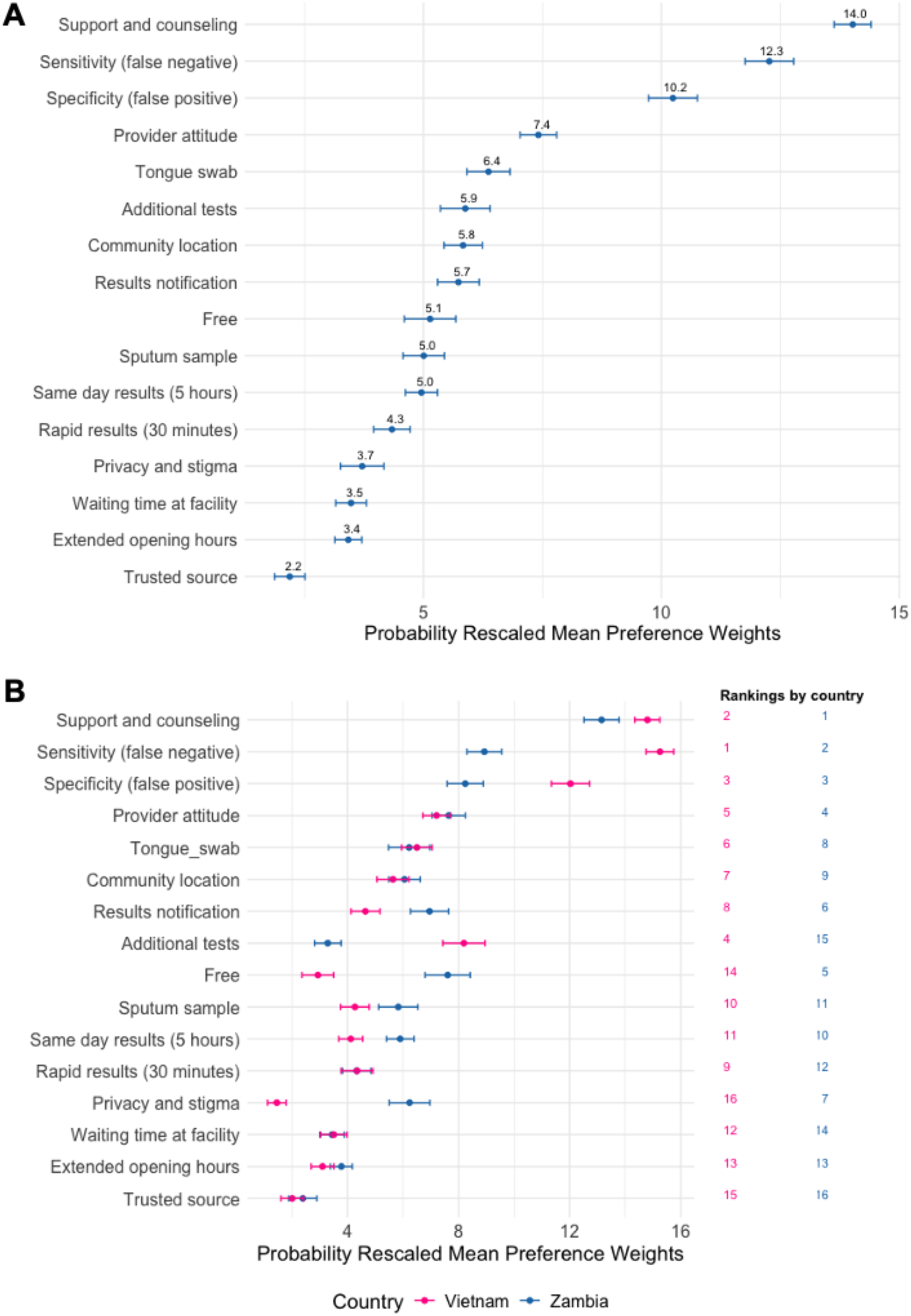
Probability rescaled average preference weights (n=356) for the overall cohort (A) and by country (B).

Country-specific estimates revealed that participants in Zambia had stronger preferences for free services (MPW=7.5 [95%CI: 6.8-8.4] vs. 3.0 [95%CI: 2.4-3.5]), a choice of results notification (MPW=6.9 [95%CI: 6.3-7.6] vs 4.6 [95%CI: 4.1-5.2), and enhanced privacy (MPW=6.2 [95%CI: 5.5-7.0] vs.1.4 [95%CI: 1.1-1.8]), while participants in Vietnam had stronger preferences for avoiding additional tests (MPW=8.3 [95%CI: 7.4-9.0] vs. 3.2 [95%CI: 2.8-3.8]) (**Figure 3B**).

Compared to older participants (30-49.9, and ≥50 years), younger participants (<30 years) placed a higher value on enhanced privacy, free services, and extended facility hours, and a lower value on TB test accuracy (**Supplementary Figure 1**). Participants who reported living with HIV had stronger preferences for test sensitivity (MPW= 13.5 [95%CI: 12.6-14.5] vs. MPW= 10.6 [95%CI: 9.8-11.4]), specificity (MPW=11.4 [95%CI: 10.5-12.5] vs MPW=9.3 [95%CI: 8.6-10.1]) and avoiding additional tests (MPW= 7.7 [95%CI: 6.5-8.8] vs MPW=4.6 [95%CI: 4.0-5.3]); those living with HIV also valued sputum samples less (MPW= 3.8 [95%CI: 3.0-4.7] vs. MPW=5.6 [95%CI: 5.0-6.3]) (**Supplementary Figure 2**). No differences in MPW were observed by sex or prior TB treatment status (**Supplementary Figures 3 and 4**).

### Heterogeneity of preferences (LCA)

Latent class analysis identified five preference groups with distinct preferences regarding TB diagnostic service features (**Figure 4**). Support and counselling were highly valued across all groups, while extended opening hours for facility-based testing and recommendations from a trusted source were consistently less valued features. Participants in Group 1 (“Supportive, Single-Time, and Accurate;” n=113, 31.7%) valued support and counseling more than any other group and strongly valued high test accuracy, as well as not having to return for additional tests. Group 2 (“Quick, Accurate, and Less-Invasive;” n=97, 27.3%) valued high test accuracy, tongue-swab-based tests, and quick turnaround times, including same-day results. Participants in Group 3 (“Service-oriented, Convenient, and Less-Invasive;” n=61, 17.1%) strongly valued tongue-swab-based testing and several service delivery features, including provider attitude, free services, enhanced privacy, and community-based testing; this group notably differed from the other four by placing relatively low value on high test accuracy. Members of Group 4 (“Free, Friendly, and Accurate;” n=46, 12.9%) placed a high value on free services and provider attitude, as well as high test accuracy; while those in Group 5 (“Result choice, Sputum Preference, and Accurate;” n=39, 11%) valued having a choice of how to receive test results, sputum-based tests, and test accuracy.

**Figure 4.**
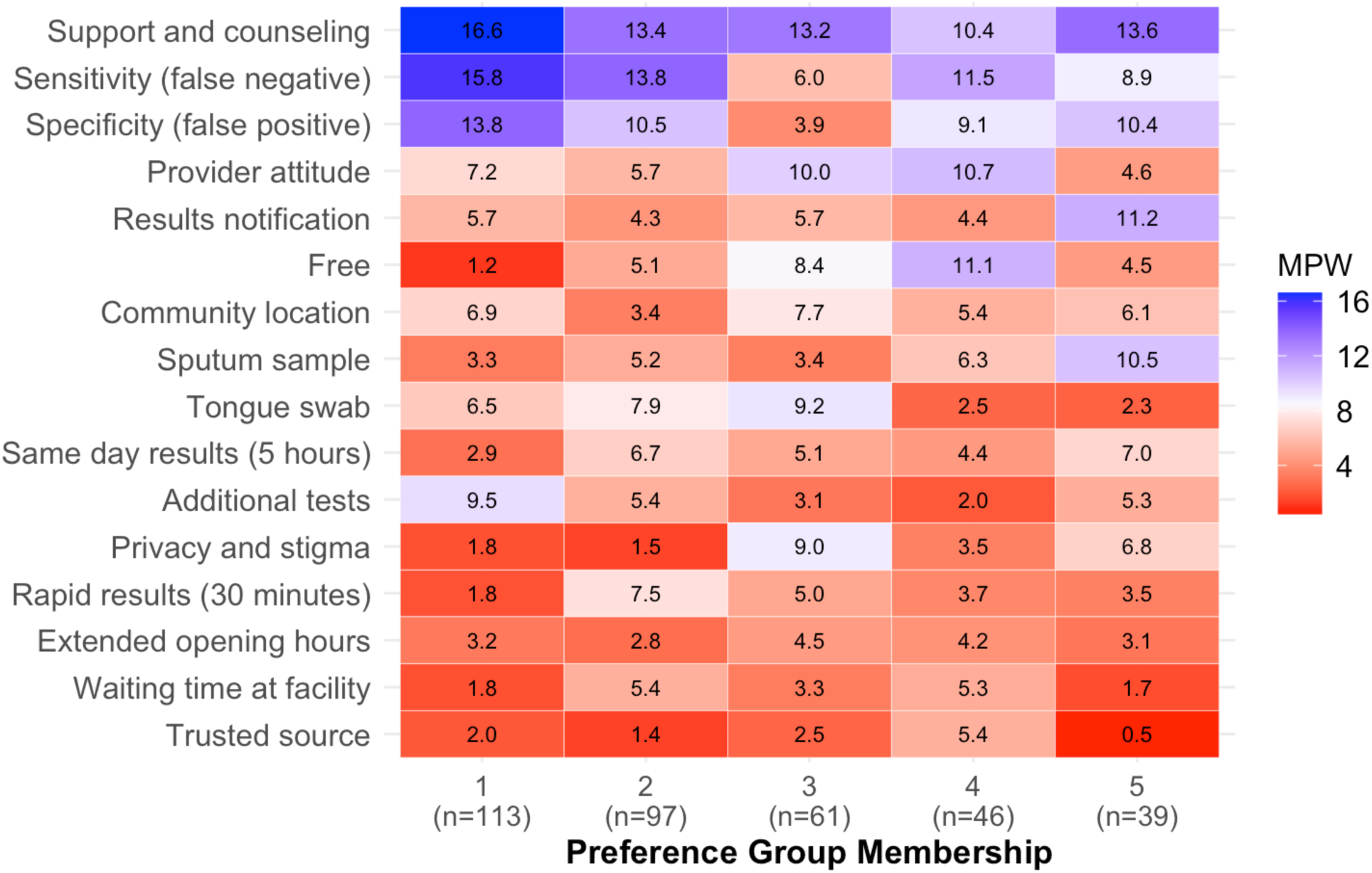
Mean preference weights for TB testing features according to latent class preference group. MPW: mean preference weights.

Participant characteristics by latent class are shown in **Supplementary Table 3**. Persons from Vietnam were substantially more likely to belong to Groups 1 and 3, while participants in Zambia were more likely to belong to Groups 2, 4, and 5. Groups differed with respect to age, education, employment status, and HIV status; however, no differences in group membership were observed by sex, prior TB testing or treatment status, or self-reported diabetes status.

## Discussion

Our study measured people’s preferences for sample type and other service features related to TB testing in two high-burden countries. Most participants preferred tongue swabs over sputum samples when asked directly. However, while tongue swab-based testing was a valued feature and preferred over sputum-based testing, the relative importance of sample type was lower compared to other TB care features such as counseling and support, test accuracy, and provider attitude.

Further, the latent class analysis identified five distinct preference groups, all of whom placed high value on support and counseling. These data show that while the development and implementation of non-sputum-based tests, including tongue swabs, are aligned with the preferences of people affected by TB, a single approach may not meet the needs of all individuals and that novel tests should be coupled with a broader focus on comprehensive TB care models that prioritize person-centered care, including robust support and counseling services and compassionate provider interactions.

Regarding sample type, while most (74.4%) participants found tongue swabs easier to provide and experienced lower discomfort and higher satisfaction with tongue swab collection, only 58.1% preferred swabs over sputum, indicating that preferences are influenced by other factors beyond ease of sample production. Differences in the perceived trustworthiness of a non-sputum-based test have been reported for urine LAM, due to how people conceptualize the association between the etiology of TB and the source of the sample [24]. In the urine LAM study, some participants questioned the plausibility that TB, a disease primarily affecting the chest, could be detected in urine [24]. Further, perceptions about accuracy and familiarity with sputum-based testing, which has been the standard sample for TB diagnosis for more than 100 years, may also influence preferences for sputum-based testing. These considerations may in part explain the differences in our results and a multi-country Discrete Choice Experiment (DCE) where participants had a small preference for sputum-based testing, although this varied by country and may also be partly attributed to most patients’ lack of experience with tongue swab-based TB testing [25].

In our study, sample type was of less relative importance than other TB testing and care delivery features. These findings align with a recent multi-country DCE, which showed that sample type was the least preferred feature of a TB test compared to other aspects like accuracy, cost, time to results, and location of testing [25]. Notably, the present study extends upon this study by quantifying the value of features of the entire TB testing and care delivery experience rather than only the TB test itself. Supportive, respectful interactions with providers, including clear communication and counseling before and after testing, were highly valued in Vietnam and Zambia, and by all five latent class groups. This highlights the critical role of person-centered care in TB diagnostic services, which is highly aligned with the END TB strategy [16]. Attention, respect, and effective communication from providers have been shown to influence care-seeking and retention throughout the TB care cascade [26–28] and extend to other health conditions across LMICs, where negative healthcare experiences are often reported [15]. Respectful care, which encompasses valuing individuals’ privacy, confidentiality, dignity, and demonstrating a compassionate attitude [6], aligns with our findings and emphasizes the importance of person-centered approaches in facilitating care-seeking and treatment adherence.

Preferences for additional TB testing service features varied considerably within and between countries. Latent Class Analysis revealed five distinct groups with differing priorities for features such as same-day results, having a choice of how results are received, enhanced privacy, free cost, and not having to return for additional testing. While four of the five groups prioritized accuracy as a key feature, one group (Group 3, “Service-oriented, Convenient, and Less-Invasive”) accounting for 17% of participants placed a much lower emphasis on high test accuracy, instead valuing aspects such as tongue-swab-based testing, provider attitude, free services, enhanced privacy, and community-based testing. This finding challenges the assumption that accuracy is universally the most prioritized feature and reflects a small subgroup potentially willing to trade lower accuracy for convenience of testing and service; it emphasizes the need for novel tests that are convenient, affordable, and easy to collect, even if they have lower accuracy. Preferences also differed to some extent by demographic factors. For instance, HIV-positive individuals prioritized accuracy and minimized additional testing, while young adults valued privacy, free services, and extended hours. Collectively, this heterogeneity highlights the importance of tailoring TB diagnostic tests and services to better appeal to and effectively reach diverse populations, which is crucial for maximizing TB diagnosis and promoting health equity.

Our study has several strengths. It was conducted in two diverse high-burden countries, enhancing the generalizability of the findings, particularly for commonly preferred and non-preferred features. Additionally, the use of standardized procedures in both countries ensured consistency and comparability, while BWS refinements through discussions with country partners and piloting contributed to local relevance and high comprehension. Furthermore, the study maintained high data quality, with less than 13% of data excluded due to low quality. Finally, having all participants experience both sample types helped minimize the hypothetical bias often found in choice experiments [29]. However, the study also had some limitations. Data collection was health center-based, excluding individuals who did not seek care, could not access primary health centers, or did not meet local TB testing requirements. Understanding the preferences of such persons is crucial as they represent those being missed by current TB services and should be prioritized in future research [30]. They may also benefit more from alternative sampling strategies since sputum collection in community settings may be challenging due to, among other factors, stigma, inability to produce and, issues handling the sample in field settings [31, 32]. Some subgroups, such as young adults and teenagers, were likely underpowered to detect differences. Also, the BWS used text descriptions without graphics, which may have led to some misunderstanding or misinterpretation despite extensive efforts by the team to explain the features and choice tasks during data collection; nevertheless, only 17% self-reported the BWS to be difficult. Lastly, the self-reporting of HIV and diabetes status may introduce minor biases likely due to underreporting, as individuals may not be aware of their HIV or diabetes status or may choose not to disclose it due to stigma.

In conclusion, participants undergoing TB testing in Vietnam and Zambia showed a clear preference for tongue swab-based testing over sputum-based testing. While tongue swab-based testing was a valued feature, the sample type was less important compared to other TB care features, including support and counseling, test accuracy, and provider attitude, which were consistently more highly valued. The latent class analysis revealed diverse preferences, including a group prioritizing free costs and convenience over test accuracy, supporting the development of new tests that improve accessibility and population coverage even if they have lower accuracy. Our findings suggest that incorporating the preferences of people affected by TB into TB care models can better meet their needs and wants, ultimately improving diagnostic outcomes and satisfaction.

## Data Availability

All data produced in the present work are contained in the manuscript

## Supplementary tables

**S1 Table.** Baseline characteristics of study participants per inclusion/exclusion of the main analysis based on data quality

| Characteristic | Included<br>n=356 | Excluded<br>n=53 | *p-value* |
| --- | --- | --- | --- |
| **Age, years, Median (IQR)** | 39 (28.5-47) | 31 (25-41) | 0.003 |
| **Female sex, n (%)** | 216 (60.7) | 32 (60.4) | 1 |
| **Country, n (%)** |  |  |  |
| Zambia | 168 (47.2) | 43 (81.1) | <0.001 |
| Vietnam | 188 (52.8) | 10 (18.9) |  |
| **Education, n (%)** |  |  |  |
| Never attended school | 8 (2.2) | 1 (1.9) | 0.09 |
| Primary school | 63 (17.7) | 16 (30.2) |  |
| Secondary school | 188 (52.8) | 28 (52.8) |  |
| Higher level | 97 (27.2) | 8 (15.1) |  |
| **Employment, n (%)** |  |  |  |
| Not employed | 75 (21.1) | 18 (34.0) | 0.02 |
| Yes, informal work | 68 (19.1) | 16 (30.2) |  |
| Yes, office job | 58 (16.3) | 4 (7.5) |  |
| Yes, self employed | 109 (30.6) | 13 (24.5) |  |
| Other | 27 (7.6) | 0 (0.0) |  |
| Student | 19 (5.3) | 2 (3.8) |  |
| **Prior testing for TB, n (%)** |  |  |  |
| Yes | 162 (45.5) | 19 (35.8) | 0.25 |
| No | 189 (53.1) | 34 (64.2) |  |
| Not sure | 5 (1.4) | 0 (0.0) |  |
| **Prior TB treatment, n (%)** |  |  |  |
| Yes | 51 (14.3) | 2 (3.8) | 0.06 |
| No |  |  |  |
| **HIV status <sup>\*\*</sup>, n (%)** |  |  |  |
| Positive | 89 (25.0) | 9 (17.0) | 0.004 |
| Negative | 168 (47.2) | 37 (69.8) |  |
| Not sure | 98 (27.5) | 6 (11.3) |  |
| **Diabetes<sup>\*</sup>, n (%)** |  |  |  |
| Yes | 8 (2.2) | 0 (0.0) | 0.37 |
| No | 343 (96.3) | 53 (100.0) |  |
| Not sure | 5 (1.4) | 0 (0.0) |  |

\* Self-reported, # two people declined to answer

**S2 Table.** Anchored mean preference weights using a Zero-Anchored Interval Scale (n=356)

| **Feature** | **Mean** | **Lower 95% CI** | **Upper 95% CI** |
| --- | --- | --- | --- |
| Tongue swab | 48.83 | 45.57 | 52.09 |
| Sputum sample | 40.80 | 36.64 | 44.97 |
| Sensitivity (false negative) | 70.65 | 67.95 | 73.35 |
| Specificity (false positive) | 61.41 | 58.35 | 64.47 |
| Additional tests | 44.43 | 41.13 | 47.74 |
| Rapid results (30 minutes) | 40.86 | 37.16 | 44.56 |
| Same day results (5 hours) | 43.87 | 40.15 | 47.60 |
| Free | 39.27 | 34.50 | 44.04 |
| Provider attitude | 54.6 | 51.51 | 57.69 |
| Waiting time at facility | 36.01 | 32.07 | 39.95 |
| Extended opening hours | 36.98 | 33.37 | 40.58 |
| Community location | 45.97 | 42.38 | 49.57 |
| Support and counseling | 76.76 | 74.51 | 79.01 |
| Privacy and stigma | 32.62 | 28.19 | 37.06 |
| Trusted source | 25.73 | 21.95 | 29.51 |
| Results notification | 44.70 | 40.91 | 48.50 |
| **Anchor** | **0.00** | **\*** | **\*** |

**S3 table.** Baseline characteristics by LCA membership group

| Variable | Preference Group Membership |  |  |  |  | *p-value* |
| --- | --- | --- | --- | --- | --- | --- |
|  | 1 | 2 | 3 | 4 | 5 |  |
| n= | 113 | 97 | 61 | 46 | 39 |  |
| **Country, n (%)** |  |  |  |  |  |  |
| Zambia | 19 (16.8) | 31 (32.0) | 49 (80.3) | 34 (73.9) | 35 (89.7) | <0.001 |
| Vietnam | 94 (83.2) | 66 (68.0) | 12 (19.7) | 12 (26.1) | 4 (10.3) |  |
| **Age, Median (IQR)** | 42 (33-48) | 41 (32-50) | 34 (25-40) | 39 (30-47) | 31 (25-46) | 0.002 |
| **Female sex, n (%)** | 66 (58.4) | 62 (63.9) | 37 (60.7) | 26 (56.5) | 25 (64.1) | 0.88 |
| **Education, n (%)** |  |  |  |  |  |  |
| Never attended school | 0 (0.0) | 0 (0.0) | 5 (8.2) | 2 (4.3) | 1 (2.6) | <0.001 |
| Primary school | 8 (7.1) | 10 (10.3) | 9 (14.8) | 18 (39.1) | 18 (46.2) |  |
| Secondary school | 65 (57.5) | 54 (55.7) | 34 (55.7) | 19 (41.3) | 16 (41.0) |  |
| Higher level | 40 (35.4) | 33 (34.0) | 13 (21.3) | 7 (15.2) | 4 (10.3) |  |
| **Employment, n (%)** |  |  |  |  |  |  |
| Not employed | 12 (10.6) | 24 (24.7) | 13 (21.3) | 18 (39.1) | 8 (20.5) | 0.002 |
| Yes, informal work | 19 (16.8) | 18 (18.6) | 11 (18.0) | 10 (21.7) | 10 (25.6) |  |
| Yes, office job | 20 (17.7) | 16 (16.5) | 9 (14.8) | 4 (8.7) | 9 (23.1) |  |
| Yes, self employed | 47 (41.6) | 19 (19.6) | 20 (32.8) | 12 (26.1) | 11 (28.2) |  |
| Other | 10 (8.8) | 14 (14.4) | 2 (3.3) | 1 (2.2) | 0 (0.0) |  |
| Student | 5 (4.4) | 6 (6.2) | 6 (9.8) | 1 (2.2) | 1 (2.6) |  |
| **Prior testing for TB, n (%)** |  |  |  |  |  |  |
| Yes | 57 (50.4) | 45 (46.4) | 23 (37.7) | 20 (43.5) | 17 (43.6) | 0.62 |
| No | 55 (48.7) | 49 (50.5) | 37 (60.7) | 26 (56.5) | 22 (56.4) |  |
| Not sure | 1 (0.9) | 3 (3.1) | 1 (1.6) | 0 (0.0) | 0 (0.0) |  |
| **Prior TB treatment, n (%)** |  |  |  |  |  |  |
| Yes | 19 (16.8) | 13 (13.4) | 8 (13.1) | 7 (15.2) | 4 (10.3) | 0.87 |
| No | 94 (83.2) | 84 (86.6) | 53 (86.9) | 39 (84.8) | 35 (89.7) |  |
| **HIV status<sup>\*\*</sup>, n (%)** |  |  |  |  |  |  |
| Positive | 33 (29.2) | 23 (23.7) | 14 (23.0) | 12 (26.1) | 7 (17.9) | <0.001 |
| Negative | 43 (38.1) | 34 (35.1) | 37 (60.7) | 23 (50.0) | 31 (79.5) |  |
| Not sure | 37 (32.7) | 40 (41.2) | 10 (16.4) | 10 (21.7) | 1 (2.6) |  |
| **Diabetes, n (%)** |  |  |  |  |  |  |
| Yes | 5 (4.4) | 1 (1.0) | 2 (3.3) | 0 (0.0) | 0 (0.0) | 0.53 |
| No | 106 (93.8) | 94 (96.9) | 58 (95.1) | 46 (100.0) | 39 (100.0) |  |
| Not sure | 2 (1.8) | 2 (2.1) | 1 (1.6) | 0 (0.0) | 0 (0.0) |  |

## Supplementary figures

**S1 Figure.**
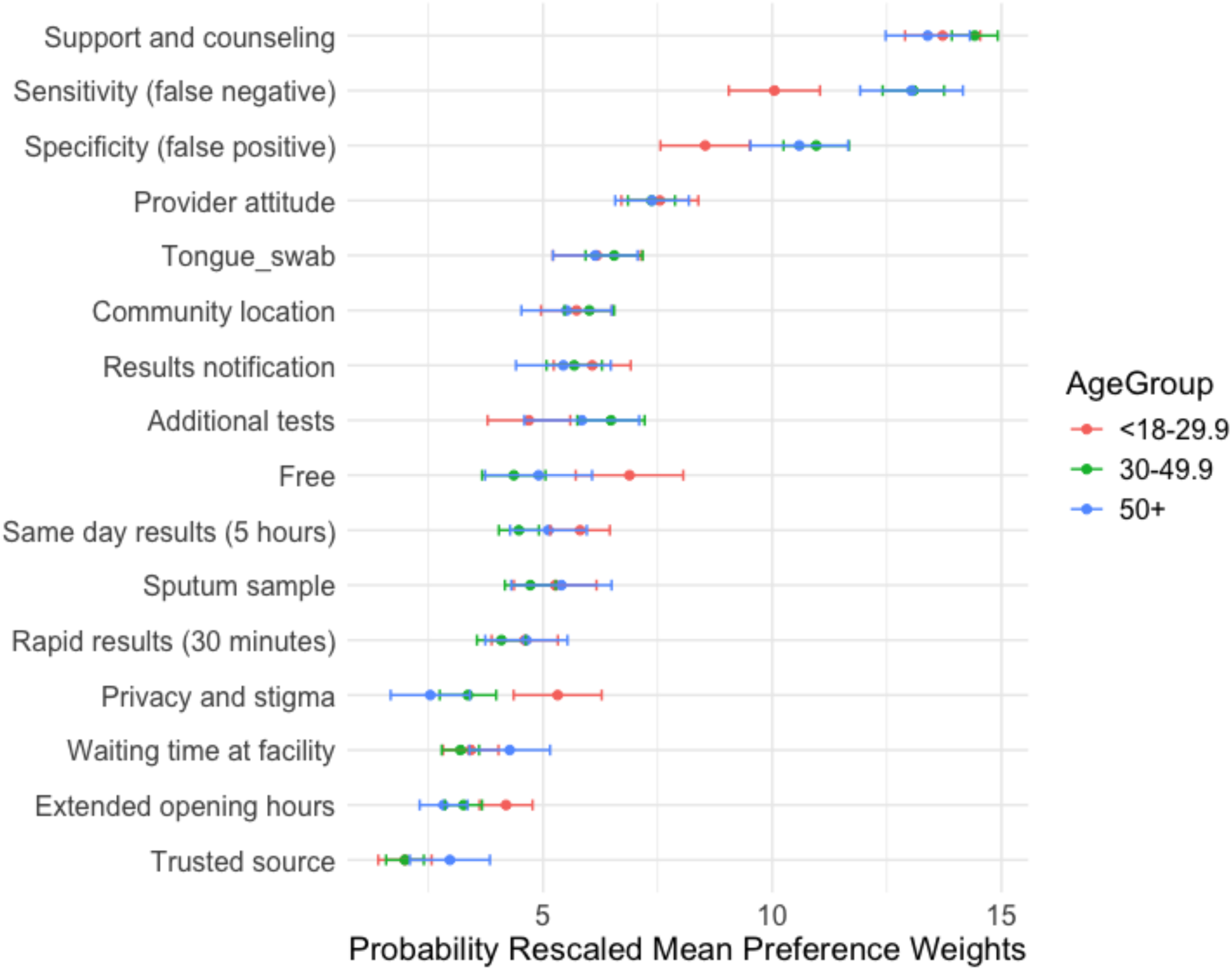
Mean preference weights by age group

**S2 Figure.**
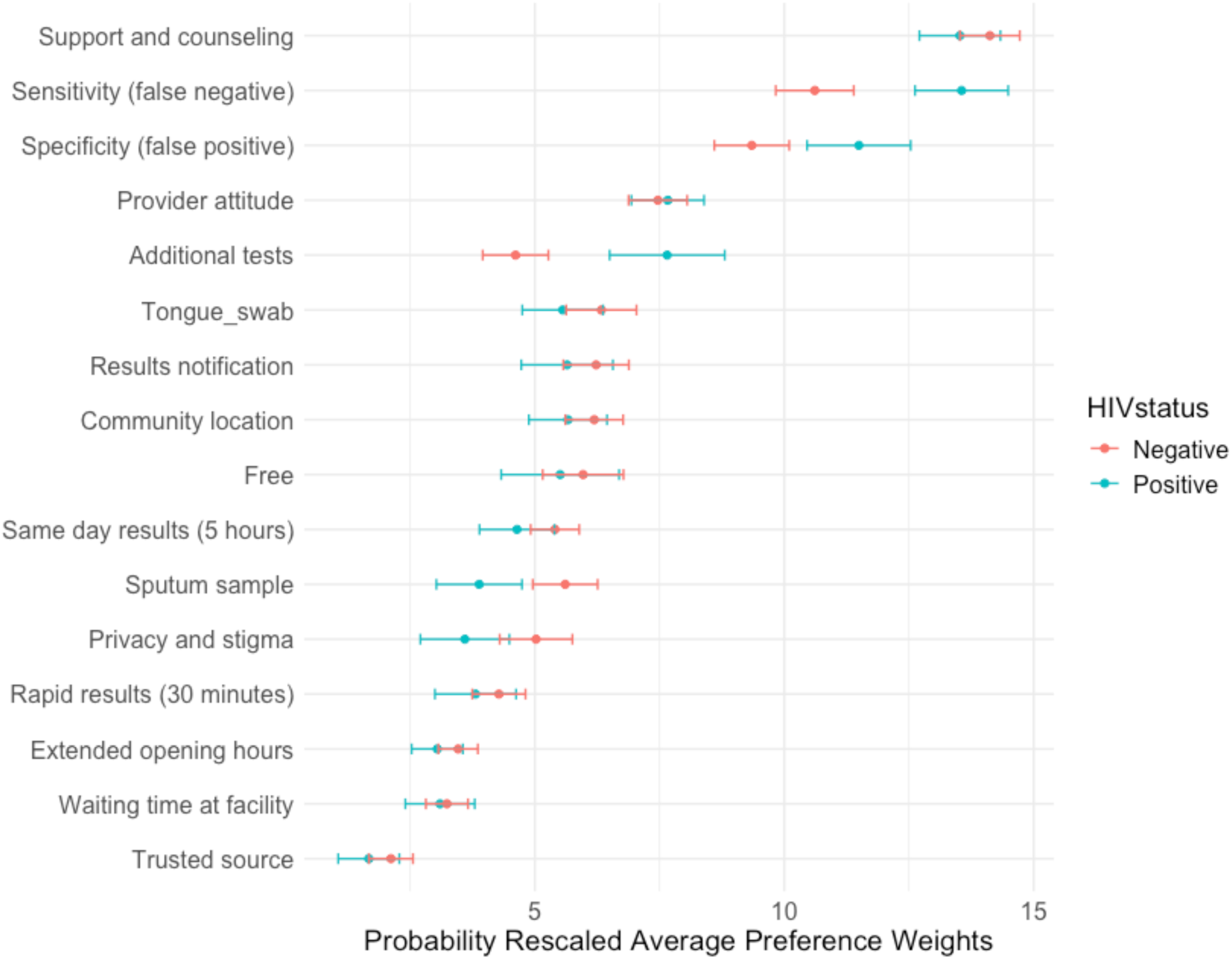
Mean preference weights by HIV status (n=257)

**S3 Figure.**
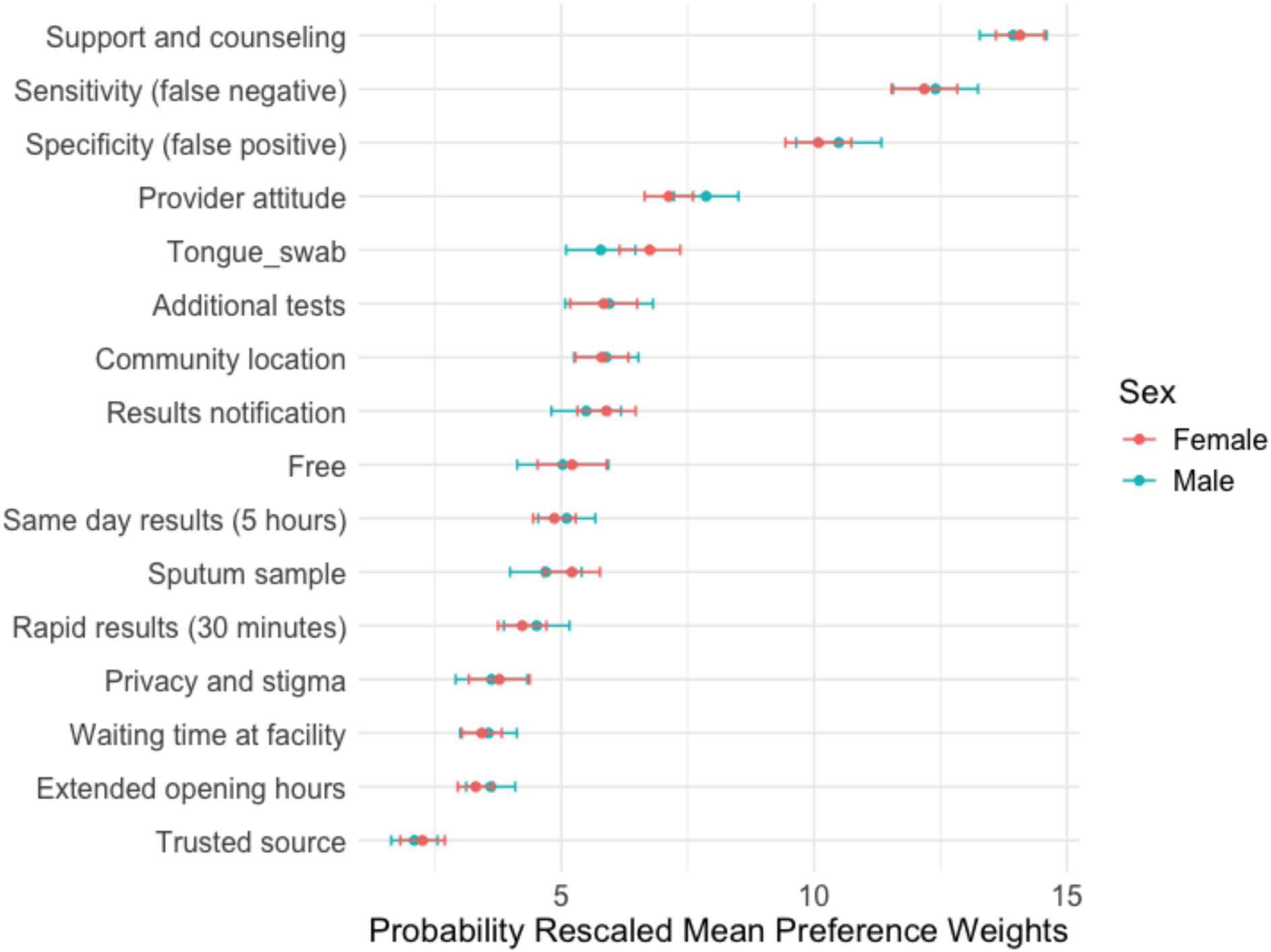
Mean preference weights by sex (n=356)

**S4 Figure.**
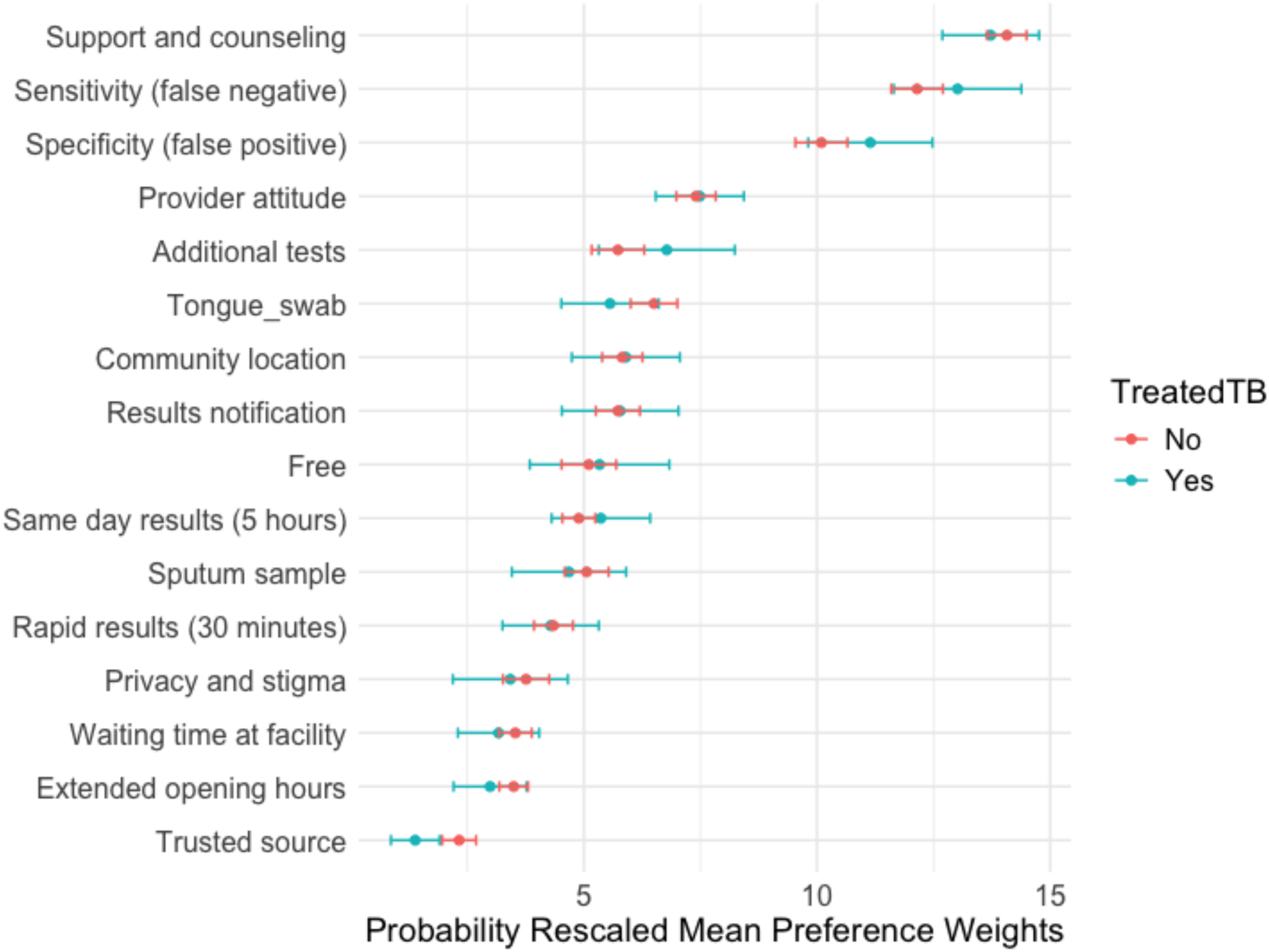
Mean preference weights by prior TB treatment status (n= 356)

